# Single-cell transcriptome-wide Mendelian randomization and colocalization analyses reveal immune-cell-specific mechanisms and actionable drug targets in prostate cancer

**DOI:** 10.64898/2026.03.10.26348013

**Authors:** Yanggang Hong, Yi Wang, Yirong Wang, Feng Chen, Jiajun Li

## Abstract

Prostate cancer remains a major health burden with limited success in immune-targeted therapies. To identify immune-cell-resolved genetically prioritized targets, we integrated single-cell cis-eQTL data across 14 immune cell types, bulk eQTLs, GWAS summary statistics from PRACTICAL and FinnGen, and single-cell RNA-seq data from prostate tumors. Using Mendelian randomization and Bayesian colocalization, we prioritized 80 eGene–cell-type associations supported by both genetic association and shared-signal evidence, with prominent representation in CD4^+^ and CD8^+^ T-cell subsets. Functional analyses revealed enrichment in immune-related pathways such as antigen processing and cytokine signaling. Meta-analysis identified 52 nominally significant eGene–cell-type associations involving 25 unique genes. Single-cell transcriptomics further mapped the expression of prioritized genes, including *HLA-DQA2*, *TXN*, and *COX6B1*, across immune populations within the prostate tumor microenvironment. Drug-target mapping highlighted candidate pharmacological connections involving genes such as *IGF1R* and *FAAH*, with known drug interactions mapped via DrugBank and STRING. Our integrative framework identifies immune-cell-resolved genetic associations with prostate cancer susceptibility and prioritizes candidate targets for further functional and pharmacological investigation. These findings provide a high-resolution resource for studying genetically informed tumor–immune mechanisms and therapeutic hypotheses in prostate cancer.

## 1. Introduction

Prostate cancer is one of the most common malignancies among men worldwide and a leading cause of cancer-related mortality (Schafer et al., 2025). Its incidence increases significantly with age, particularly in individuals over 50 years old (Kratzer et al., 2025). Despite advances in early detection and treatment, a subset of patients still progress to aggressive or treatment-resistant disease, underscoring the need for deeper understanding of its molecular underpinnings and novel therapeutic approaches.

Numerous studies have sought to uncover the molecular drivers of prostate cancer, identifying alterations in androgen receptor signaling, DNA repair pathways, and cell cycle regulators as critical factors (Burdak-Rothkamm et al., 2020; Dai et al., 2023; He et al., 2022). These insights have informed the development of targeted therapies, including androgen deprivation therapy (ADT) and next-generation antiandrogens (Chakrabarti et al., 2025; Gillessen et al., 2025). However, resistance often emerges, and the effectiveness of current drugs remains limited, prompting continued efforts to identify new biomarkers and therapeutic targets (Cai et al., 2023; Swami et al., 2020).

Emerging evidence suggests that the immune system plays a complex and context-dependent role in prostate cancer (Chen et al., 2024; Jiang et al., 2025). Although traditionally considered poorly immunogenic, prostate tumors contain diverse immune-cell populations within their microenvironment (Chen et al., 2024). Therapeutic strategies to enhance antitumor immunity, including T-cell engagers and combinations of irreversible electroporation with oncolytic viruses, have been reviewed or proposed (Hage Chehade et al., 2025; Xia et al., 2025). These immune components may contribute to tumor progression, immune evasion, or therapeutic resistance. Immunotherapies such as immune checkpoint inhibitors have shown limited efficacy in prostate cancer, highlighting a crucial gap in understanding the immune regulatory mechanisms specific to this disease (Sridaran et al., 2023). Therefore, there is an urgent need to systematically investigate immune-related genetic drivers of prostate cancer and their downstream pathways. Identifying immune-cell-resolved genetically prioritized genes could provide mechanistic insights into the interplay between host immunity and prostate cancer susceptibility and nominate candidates for subsequent functional investigation.

Human genetic studies can inform drug-target prioritization and drug development (Carss et al., 2023; Reay & Cairns, 2021). In prostate cancer, genome-wide association studies (GWAS) have identified hundreds of susceptibility loci, shedding light on inherited genetic variation contributing to disease risk (Akamatsu et al., 2012; Farashi et al., 2019; Schumacher et al., 2018). However, most GWAS signals reside in non-coding regions, limiting biological interpretation. To bridge this gap, transcriptome-wide association studies (TWAS) and Mendelian randomization (MR) approaches have been used to integrate genetically predicted gene expression with disease GWAS and prioritize susceptibility-associated genes (Reay & Cairns, 2021). These approaches often integrate GWAS data with expression quantitative trait loci (eQTL) from bulk tissues to identify putative disease-associated genes (Porcu et al., 2019; Richardson et al., 2020). Several TWAS studies have identified candidate susceptibility genes for prostate cancer (Liu et al., 2022; Wu et al., 2019). Nevertheless, bulk eQTLs may mask cell-type-specific regulatory effects, especially in heterogeneous tissues like blood or tumors. Recently, single-cell eQTL (sc-eQTL) approaches have emerged, enabling the resolution of eQTL effects at individual immune cell types (Nathan et al., 2022; van der Wijst et al., 2018; Yazar et al., 2022). Integrating sc-eQTL data with GWAS using a single-cell-informed TWAS (sc-TWAS) design can improve causal inference resolution and uncover cell-specific regulatory mechanisms that are otherwise undetectable in bulk data (Mai et al., 2025). This strategy holds promise for identifying immune-cell-resolved genetically regulated eGenes associated with prostate cancer susceptibility.

In this study, we systematically integrated single-cell and bulk eQTL data with large-scale prostate cancer GWAS using a two-sample MR framework (***Figure 1***). We aimed to identify genetically regulated eGenes associated with prostate cancer susceptibility at immune-cell resolution, assess whether these associations were supported by shared genetic signals, validate their reproducibility across independent cohorts, characterize their biological and cellular context, and explore candidate pharmacological connections. Our approach provides novel insights into the immune-related genetic architecture of prostate cancer and offers a genetically informed resource for prioritizing candidates for subsequent functional and therapeutic investigation.

**Figure 1.**
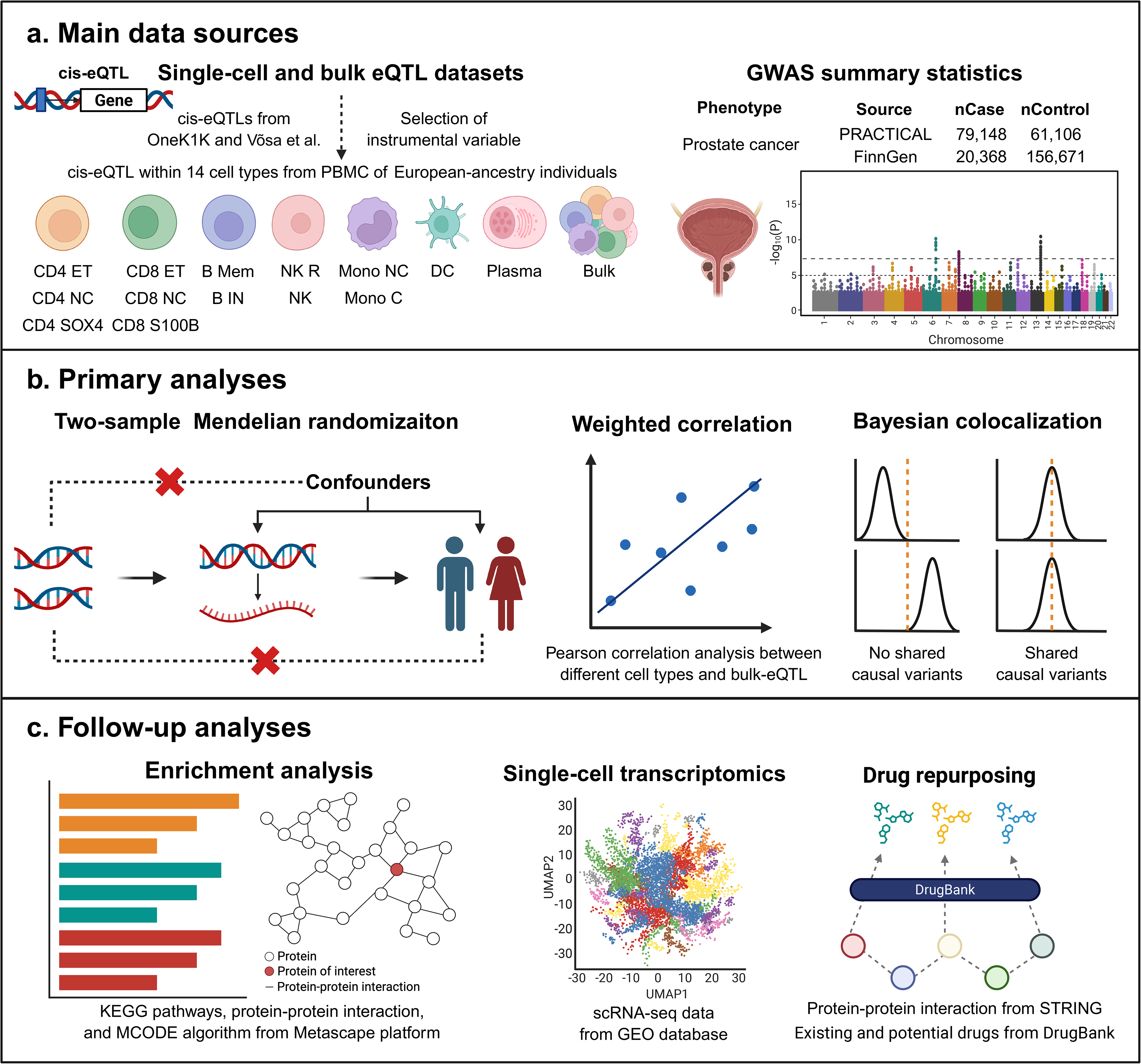
Overview of study design and workflow. (A) Main data sources: Single-cell and bulk cis-eQTL datasets from European-ancestry individuals were used to define immune-cell-resolved gene expression regulation. GWAS summary statistics for prostate cancer were obtained from the PRACTICAL and FinnGen consortia. (B) Primary analyses: Two-sample Mendelian randomization (MR) was used to infer causal effects, followed by weighted correlation analysis across immune cell types and Bayesian colocalization to assess shared genetic variants. (C) Follow-up analyses: Functional enrichment, protein–protein interaction (PPI) network analysis, single-cell RNA sequencing (scRNA-seq) mapping of prioritized eGenes within the prostate tumor microenvironment, and drug-target analysis via STRING and DrugBank were performed to characterize biological and pharmacological context.

## 2. Results

### 2.1 MR identifies genetically prioritized eGenes associated with prostate cancer susceptibility

We performed two-sample MR analyses using cis-eQTL data from 14 immune cell types and bulk blood to identify genetically predicted gene-expression effects associated with prostate cancer susceptibility (***Tables S2–3***). The final reported single-cell MR results included 413 significant gene–cell-type associations involving 243 unique genes, while the bulk-blood MR results included 64 significant associations involving 64 unique genes (FDR < 0.05). The largest numbers of significant gene–cell-type associations were observed in CD4 NC, CD8 ET, NK, and CD8 NC populations.

To assess the concordance of MR effect estimates across eQTL contexts, we calculated pairwise weighted Pearson correlations using all available MR estimates rather than restricting the analysis to significant associations (***Figure 2A***; ***Tables S4–6***). High concordance was observed between closely related T-cell subsets, including CD4 NC and CD8 NC (*r* = 0.78). Correlations between sc-eQTL and bulk-eQTL data derived MR results were generally weaker, suggesting that cell-resolved eQTL analyses capture regulatory heterogeneity that may be obscured in bulk blood eQTL analyses.

**Figure 2.**
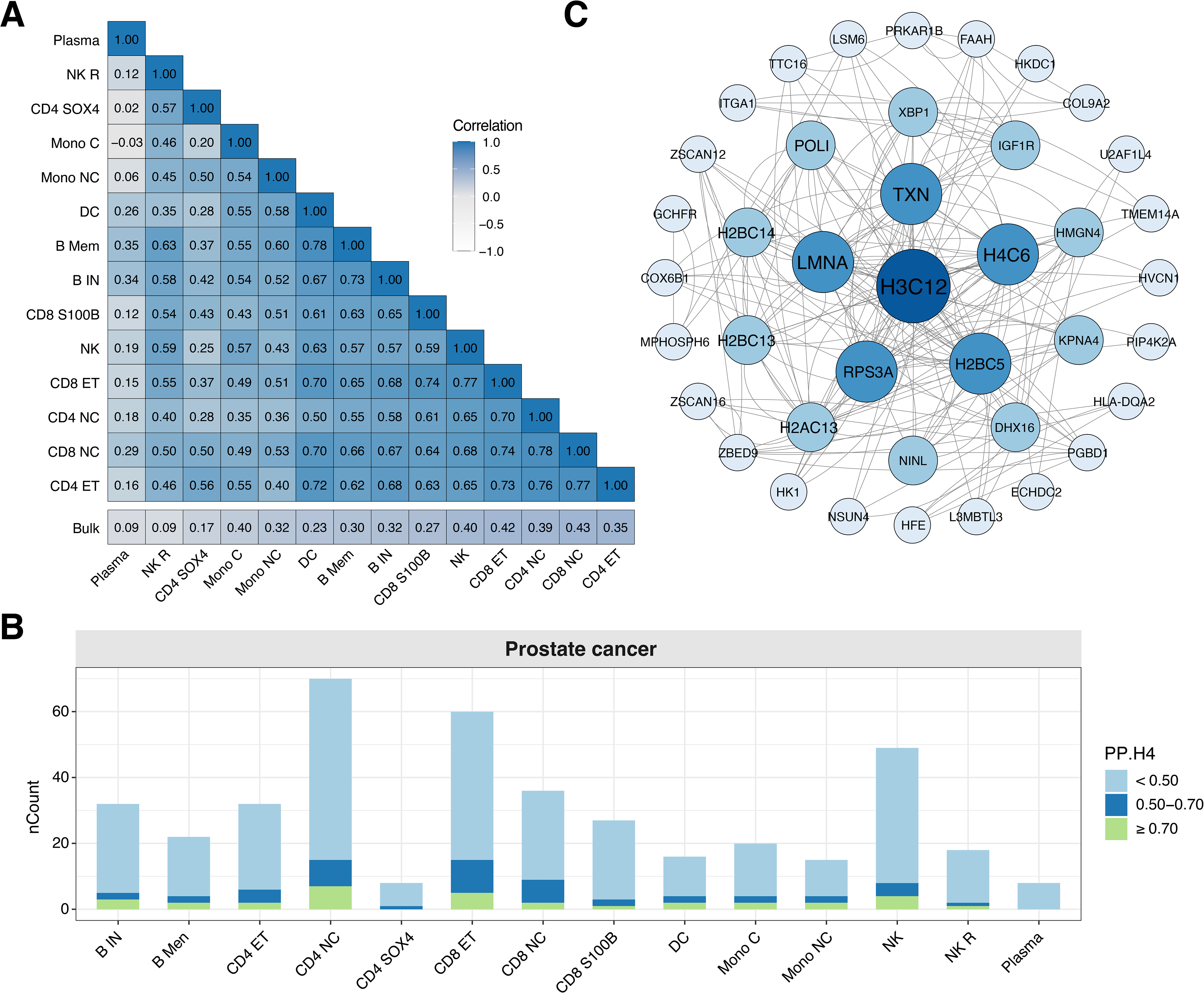
Correlation, colocalization, and protein interaction network in prostate cancer. (A) Weighted Pearson correlation matrix of MR results across immune cell types and bulk-eQTL, showing similarity in genetically predicted effect estimates. (B) Number of prostate cancer-associated eGenes in each immune cell type with varying levels of colocalization evidence. (C) PPI network of colocalized eGenes prioritized by Bayesian colocalization (PP.H4 ≥ 0.50), with node size representing connectivity.

### 2.2 Colocalization and protein network analysis of candidate prostate cancer susceptibility genes

Among the prioritized eGenes showing significant MR associations, 80 eGene–cell-type associations exhibited evidence of colocalization (PP.H4 ≥ 0.50), with 33 associations involving 19 unique genes reaching strong support (PP.H4 ≥ 0.70) (***Table S7***). The number of colocalized eGenes varied by cell type, with the highest counts observed in CD4 NC, CD8 ET, CD8 NC, and NK, suggesting heterogeneity in the genetic regulatory signals captured across immune-cell populations (***Figure 2B***). The distribution of these colocalized eGene–cell-type associations across immune-cell eQTL contexts is summarized in ***Figure S1*** and ***Table S8***. These colocalized eGenes were further mapped into the protein-protein interaction (PPI) network to explore functional interconnectivity. Network topology analysis identified highly connected nodes, including H3C12, TXN, LMNA, and H4C6, which showed relatively high degree centrality within the constructed PPI network (***Figure 2C***; ***Table S9***).

### 2.3 Functional enrichment and PPI network analysis

KEGG pathway enrichment analysis of the broader MR-prioritized candidate set identified significant enrichment in immune-related pathways, including antigen processing and presentation and Th1 and Th2 cell differentiation (***Figure 3A***; ***Table S10***). Additional enriched pathways included necroptosis and pathways related to viral infections. Metascape analysis identified enrichment in biological processes related to the regulation of lymphocyte-mediated immunity, antigen presentation, and humoral immune responses (***Table S11***).

**Figure 3.**
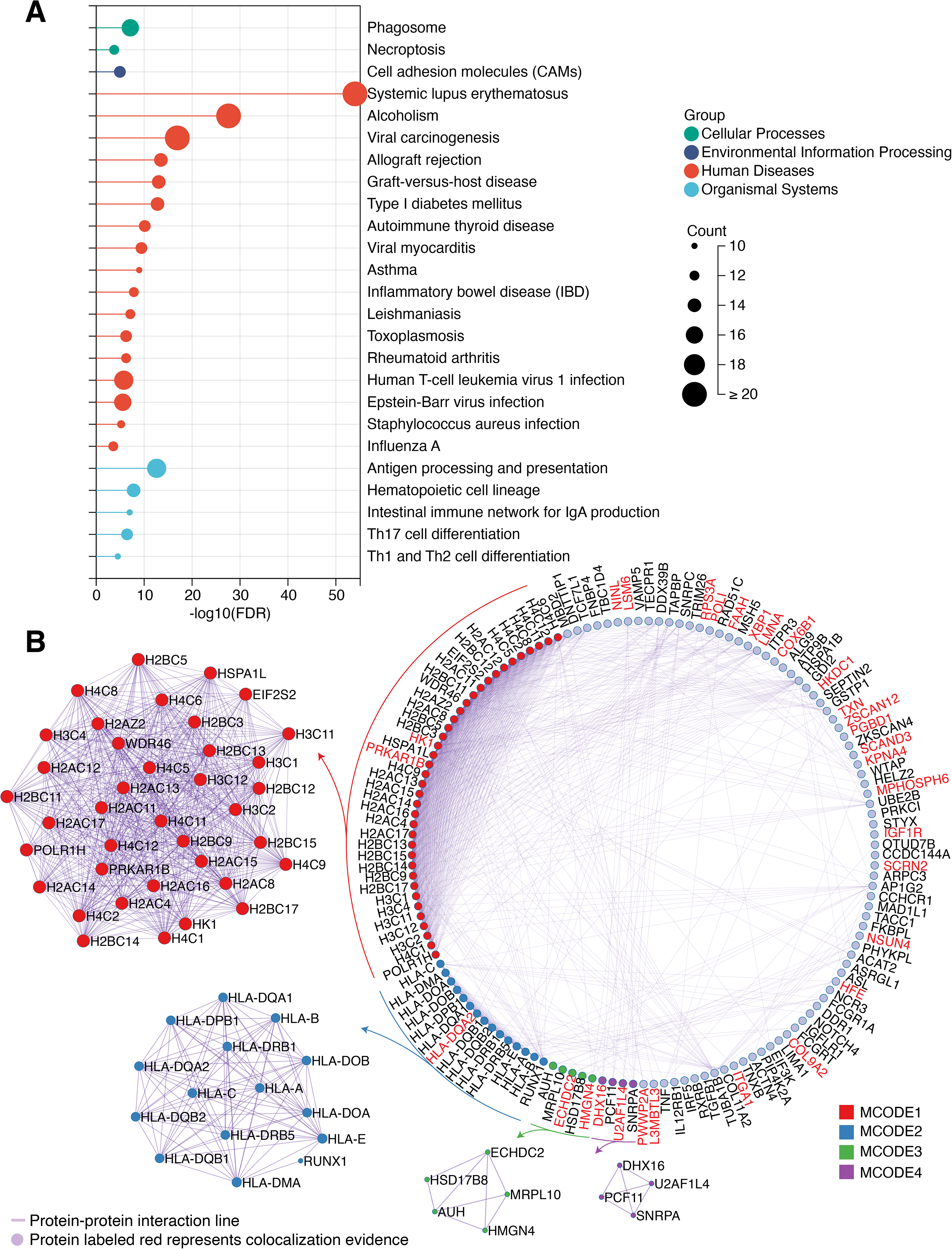
Functional enrichment and PPI module analysis of MR-prioritized prostate cancer susceptibility candidates. (A) KEGG pathway enrichment results for the broader MR-prioritized gene set. (B) PPI network and densely connected modules identified using MCODE. Red gene labels indicate candidates with colocalization support (PP.H4 ≥ 0.50) in at least one immune-cell eQTL context.

To explore the network architecture of these eGenes, we constructed a PPI network and applied the MCODE algorithm to identify densely connected modules. Four major modules (MCODE1–4) were detected (***Figure 3B***; ***Table S12***). MCODE1 predominantly comprised histone-related genes, including H2BC5, H3C12, and H2AC13, indicating that a substantial portion of the network connectivity involved chromatin- and nucleosome-associated proteins. Whether this module reflects prostate cancer-specific mechanisms, broader cellular stress responses, or age-associated regulatory processes requires further investigation. MCODE2 contained HLA class I and II genes, further implicating immune presentation mechanisms. The network architecture suggested two broad biological components: an immune-presentation module dominated by HLA-related genes and a chromatin/stress-associated component involving histone-related and broadly expressed regulatory genes.

### 2.4 Transcriptional mapping of genetically prioritized eGenes in the tumor microenvironment

To characterize the tumor-microenvironment expression context of genetically prioritized prostate cancer susceptibility-associated eGenes, we analyzed an independent prostate cancer single-cell RNA-sequencing (scRNA-seq) dataset. A total of 80 eGene–cell-type associations with colocalization evidence were examined across the major cellular compartments of the prostate tumor microenvironment, revealing heterogeneous transcriptional representation across immune, epithelial, and stromal populations. Using Seurat clustering and canonical markers, we identified 9 major cell types, including epithelial, T cells, B cells, myeloid cells, fibroblasts, and endothelial cells (***Figure 4A–C***; ***Table S13***). Marker gene expression and dot plot analysis confirmed the specificity and annotation of each cluster (***Figure 4D***). Cell composition analysis showed epithelial cells as the predominant population, with varying contributions from immune and stromal compartments across patients (***Figure 4E–F***; ***Table S14***). We first examined the complete tumor microenvironment to determine whether genetically prioritized eGenes were predominantly confined to immune compartments or also represented in epithelial and stromal populations. To further explore the potential roles of eGenes beyond immune cells, we analyzed bulk-prioritized eGenes that lacked consistent prioritization across single-cell immune cell types (***Figure 4G***). Notably, *MYO6*, *MRPS34*, and *MMAB* showed predominant expression in epithelial populations, whereas *VAMP8* showed prominent expression in myeloid cells, illustrating that genetically prioritized prostate cancer-associated genes may occupy cellular contexts beyond the immune compartments represented in the sc-eQTL analysis (***Figure 4H–K***).

**Figure 4.**
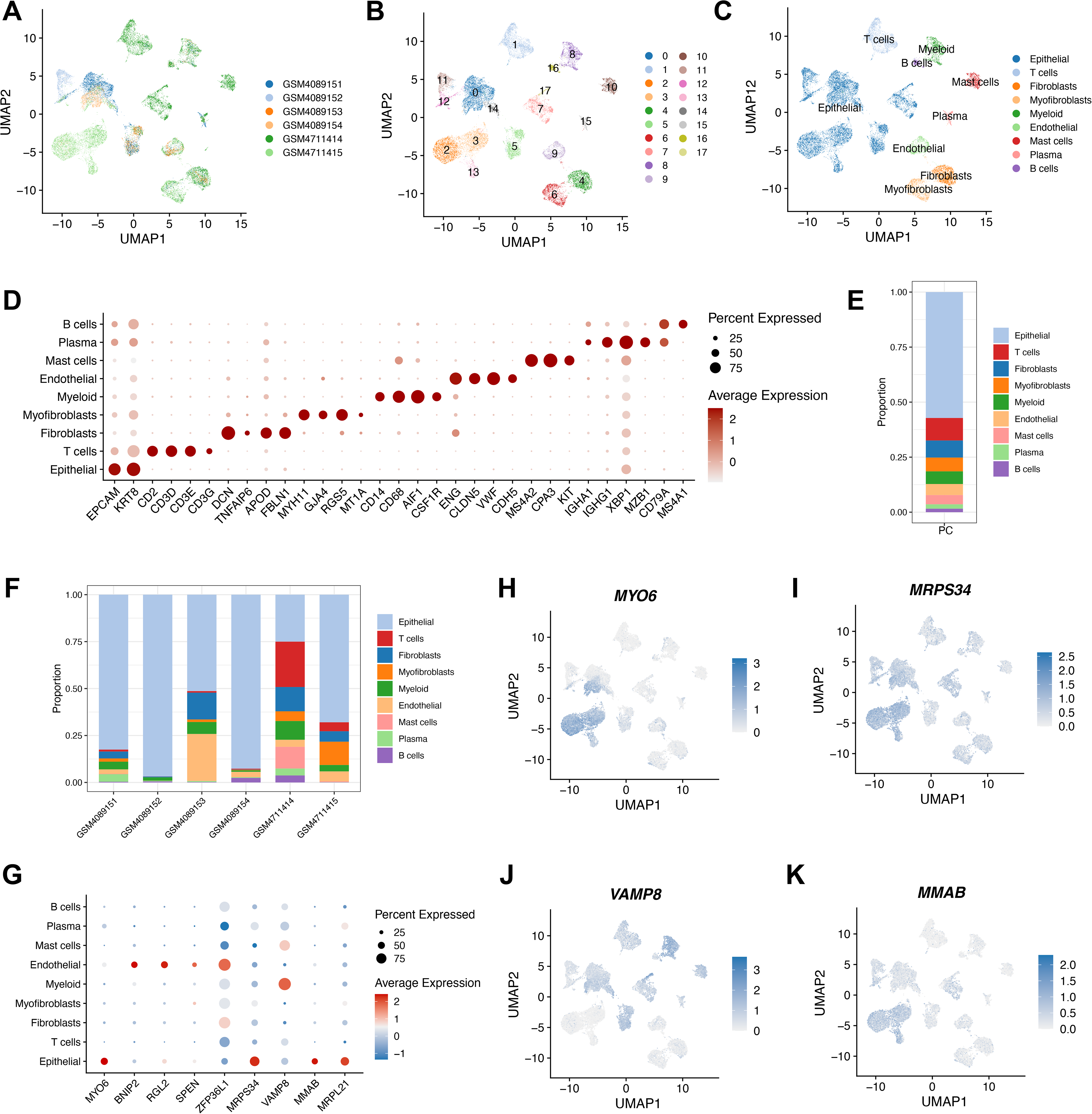
Transcriptional contextualization of genetically prioritized eGenes within the prostate tumor microenvironment. (A–C) UMAP plots showing sample identity, unsupervised clustering, and annotation of major tumor, immune, and stromal populations. (D) Dot plot of canonical markers used for cell-type annotation. (E–F) Overall cellular composition and cell-type proportions within individual patient samples. (G) Expression patterns of selected bulk-blood MR-prioritized genes across major cellular compartments. (H–K) Feature plots illustrating the expression of representative genes (*MYO6*, *MRPS34*, *VAMP8*, and *MMAB*) across annotated cell populations.

### 2.5 Expression of prioritized eGenes across lymphocyte populations

To further resolve the transcriptional distribution of genetically prioritized eGenes within lymphocyte populations, we reclustered plasma cells, T cells, and B cells from the prostate tumor microenvironment (***Figure S2A–C***). Based on canonical marker expression, we identified four major lymphocyte populations: Plasma, CD4^+^ T, CD8^+^ T, and B cells (***Figure S2D***; ***Table S15***). Expression analysis revealed heterogeneous transcriptional patterns of genetically prioritized eGenes across major lymphocyte populations (***Figure S2E***). For example, *HLA-DQA2* and *TXN* showed strong expression in B cells, whereas *TXN* was also expressed in both CD4^+^ and CD8^+^ T cells (***Figure S2F–G***). Additional eGenes such as *RPS3A*, *COX6B1*, and *XBP1* displayed heterogeneous transcriptional representation across major immune populations. These findings provide cellular expression context for the genetically prioritized associations and identify the lymphocyte populations in which selected candidates are transcriptionally represented, thereby guiding subsequent biological interpretation.

### 2.6 Cross-cohort comparison and meta-analysis of prioritized associations

To evaluate the robustness of the genetically prioritized associations, we performed replication analyses using prostate cancer GWAS summary statistics from the FinnGen consortium and integrated the results with the PRACTICAL cohort through random-effects meta-analysis. Five eGene–cell-type associations were unavailable for replication because of insufficient overlapping genetic instruments (***Table S16***). Across the two cohorts, meta-analysis identified 52 nominally significant eGene–cell-type associations involving 25 unique genes (*p* < 0.05). Several prioritized genes, including *NSUN4*, *TXN*, *HK1*, *LMNA*, and *L3MBTL3*, showed consistent effect directions between PRACTICAL and FinnGen, supporting the reproducibility of their genetically predicted associations with prostate cancer susceptibility (***Table S17***). Among these associations, 20 eGene–cell-type associations involving 11 unique genes showed strong colocalization evidence (PP.H4 ≥ 0.70) and significant meta-analysis results, with corresponding FinnGen estimates available (***Figure 5***). These included *NSUN4*, *COX6B1*, *TXN*, *PIP4K2A*, *HKDC1*, *HK*1, *LMNA*, *LSM6*, *TTC16*, *KPNA4*, and *L3MBTL3*. These associations were prioritized for further investigation based on concordant effect directions across cohorts, nominally significant meta-analysis results, and colocalization support in PRACTICAL.

**Figure 5.**
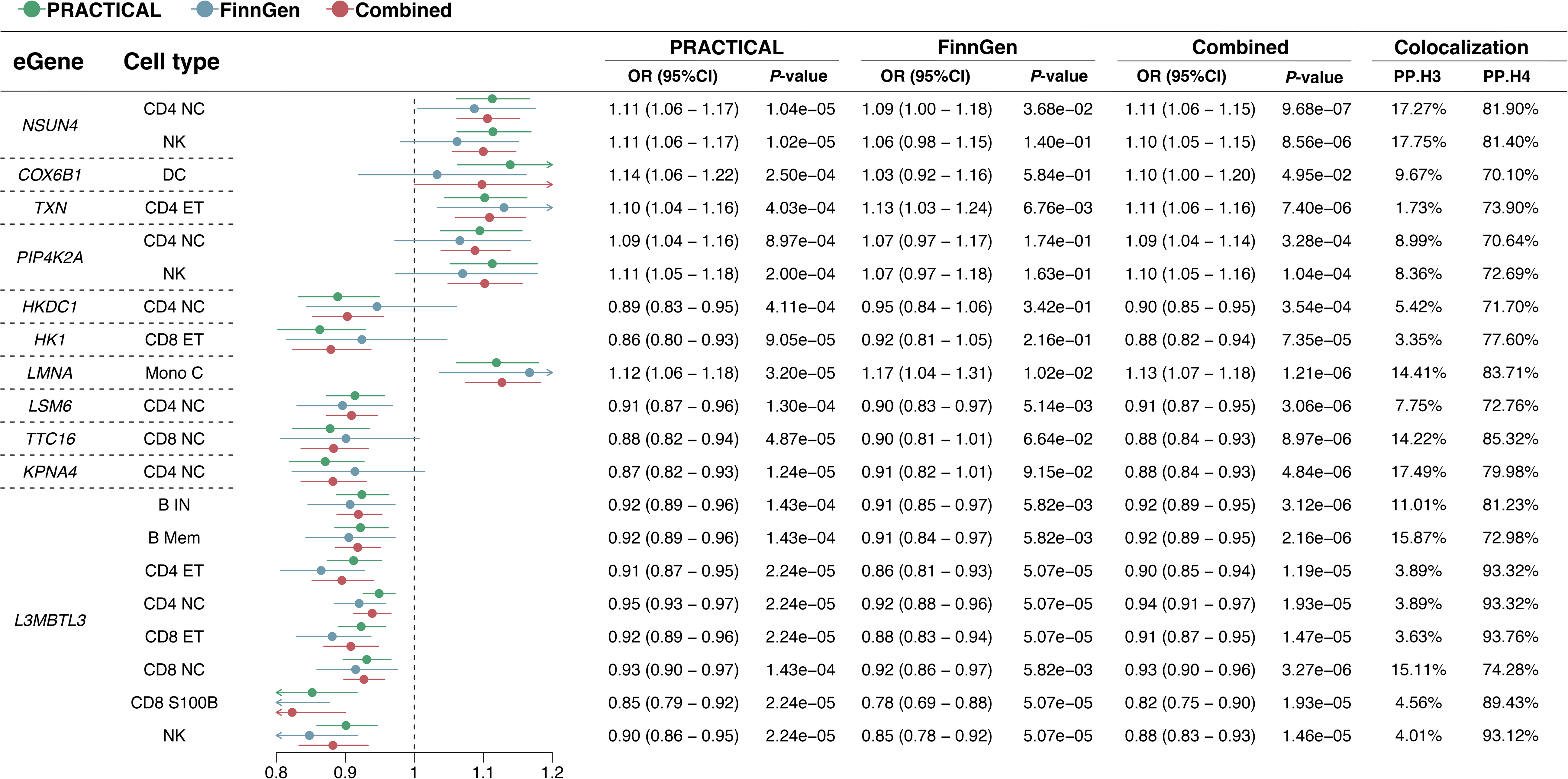
Cross-cohort effect estimates of prioritized eGene–cell-type associations. Associations with available FinnGen estimates, significant meta-analysis results (*p* < 0.05), and strong colocalization evidence in PRACTICAL (PP.H4 ≥ 0.70) are displayed. Odds ratios and 95% confidence intervals are shown for PRACTICAL, FinnGen, and combined meta-analysis.

### 2.7 Drug-target mapping identifies candidate pharmacological connections

DrugBank searches identified compound–protein associations involving genetically prioritized prostate cancer susceptibility genes (***Tables S18–19***). Separately, we compiled records of drugs and diagnostic agents associated with prostate cancer and their associated proteins (***Table S20***). Functional annotations of both protein groups indicated their involvement in biological pathways related to tumor biology and treatment response (***Tables S21–22***). Subsequently, we examined STRING associations between the two protein groups. The connected portion of the network comprised 16 genetically prioritized candidate proteins, 15 prostate cancer drug-associated proteins, 10 candidate compounds, and two prostate cancer drugs, estradiol and etoposide (***Figure 6***). Cholic acid was associated with COX6B1, while acetaminophen, cannabidiol, fostamatinib, propofol, and thiopental were associated with FAAH. Brigatinib, mecasermin, mecasermin rinfabate, and teprotumumab were linked to IGF1R. Estradiol-associated proteins included ESR1, ESR2, and CYP19A1, whereas etoposide-associated proteins included TOP2A and TOP2B.

**Figure 6.**
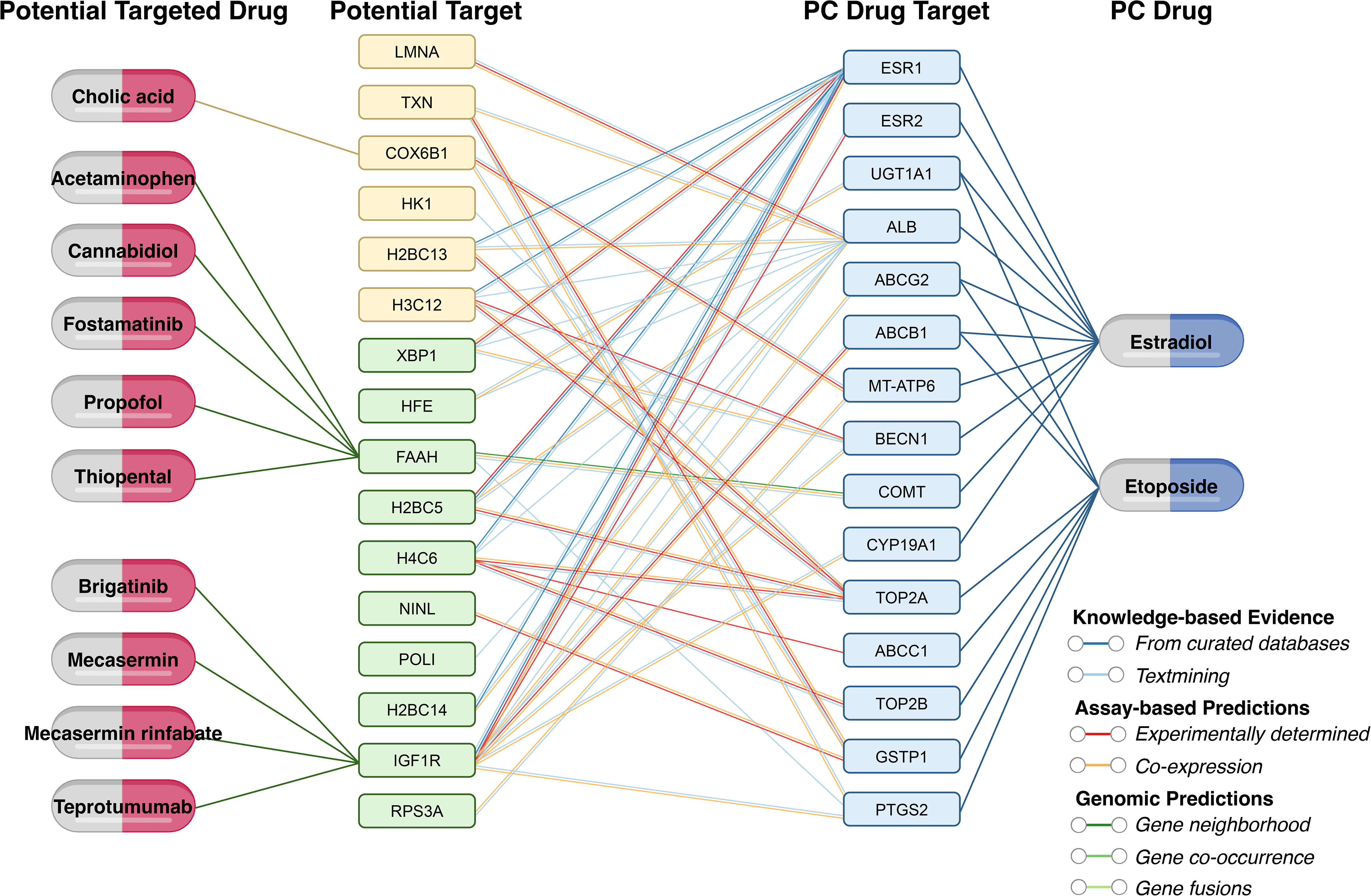
Integrated network of genetically prioritized candidate targets, drug-associated proteins, and compounds in prostate cancer. The network integrates three types of relationships: (1) associations between genetically prioritized candidate targets and compounds recorded in DrugBank; (2) associations between prostate cancer drugs and their associated proteins, including pharmacological targets, enzymes, transporters, and carriers; and (3) STRING protein associations connecting the two protein groups. Yellow nodes indicate candidate targets with strong colocalization support (PP.H4 ≥ 0.70), whereas green nodes indicate those with moderate colocalization support (0.50 ≤ PP.H4 < 0.70). STRING evidence colors apply to protein–protein associations. Only proteins linked by at least one retained STRING association between the two groups, together with their associated compounds, are displayed.

## 3. Discussion

Prostate cancer remains a complex and heterogeneous disease with limited insight into the immune-related mechanisms driving its initiation and progression (Haffner et al., 2021). In this study, we employed a comprehensive analytical framework integrating sc-TWAS, Bayesian colocalization, scRNA-seq, and drug repurposing approaches to systematically identify cell-type-resolved genetically prioritized eGenes associated with prostate cancer susceptibility. Our findings point to a complex immune-related genetic architecture of prostate cancer susceptibility, in which distinct regulatory programs emerge across different immune-cell contexts. T-cell and NK-cell populations showed particularly prominent genetically regulated associations, while prioritized genes converged on pathways related to antigen presentation, redox balance, chromatin regulation, and cellular stress. This pattern suggests that prostate cancer susceptibility may arise from the interplay between immune regulation and more broadly conserved cellular programs rather than from a single immune-cell-specific mechanism. Several signals were replicated across independent cohorts and supported by colocalization, providing prioritized candidates for further mechanistic investigation. These findings should also be considered in the context of prostate ageing. Age-associated inflammation, epigenetic remodeling, and cellular stress responses may contribute to the chromatin-, redox-, and stress-related programs identified here, suggesting shared biological processes linking immune regulation, tissue homeostasis, and prostate cancer susceptibility (Bianchi-Frias et al., 2010; Damaschke et al., 2013; Sun et al., 2026).

Several representative genetically prioritized eGenes identified in our analysis showed distinct transcriptional patterns across immune populations and provided biologically plausible candidates for further investigation. The network and enrichment analyses further suggest that the prioritized genes do not converge on a single immune pathway. Instead, they span at least two complementary biological components. One component centers on antigen processing and HLA-mediated immune presentation, whereas another involves chromatin organization, redox control, mitochondrial metabolism, and cellular stress. The latter processes are broadly conserved across cell types and may influence prostate cancer susceptibility by shaping both immune-cell function and the wider tissue microenvironment. This modular view provides a more biologically coherent interpretation than assigning mechanistic importance to individual highly connected genes solely on the basis of network centrality. *HLA-DQA2*, which encodes an HLA class II DQ α-chain, showed an inverse genetically predicted association when instrumented in B Mem cells and was transcriptionally detectable in tumor-associated B cells. These observations identify B-cell antigen-presentation biology as a plausible context for further mechanistic investigation. Enrichment results supporting negative regulation of antigen presentation and humoral immune responses further reinforce this interpretation. In contrast, *TXN* (Thioredoxin), a regulator of redox homeostasis and immune signaling, showed risk-increasing genetically predicted associations in several T-cell eQTL contexts, including CD4 NC, CD4 ET, and CD8 ET, and was expressed across multiple lymphocyte populations. Given its established roles in redox regulation and immune-cell signaling, these findings raise the hypothesis that *TXN*-related redox programs may contribute to the immune environment associated with prostate cancer susceptibility (Bradford et al., 2024; Matsuzawa, 2017). *RPS3A* showed broad lymphocyte expression, indicating a broadly distributed rather than cell-restricted transcriptional pattern. *COX6B1* encodes a subunit of cytochrome c oxidase involved in mitochondrial respiration (Abdulhag et al., 2015). In our analysis, it showed risk-increasing genetically predicted associations across several immune-cell eQTL contexts, including CD4 NC, CD4 ET, CD8 NC, CD8 ET, and B Mem. Its expression pattern, particularly within CD4^+^ T-cell populations, raises the possibility that mitochondrial metabolic programs may influence immune-cell states relevant to prostate cancer susceptibility (Buck et al., 2016; Steinert et al., 2021).

The identification of pharmacologically connected candidates among genetically prioritized prostate cancer-associated eGenes provides a framework for subsequent translational investigation. *IGF1R* signaling has been implicated in prostate cancer biology (Elemam et al., 2024). Our DrugBank analysis identified pharmacological connections between IGF1R and compounds including brigatinib and teprotumumab. *FAAH* (fatty acid amide hydrolase) participates in endocannabinoid metabolism and has also been implicated in NLRP3 stabilization (Zhu et al., 2023). Our DrugBank analysis identified associations with compounds including cannabidiol and acetaminophen. FAAH-dependent metabolism contributes to the analgesic action of acetaminophen (Dalmann et al., 2015). These findings provide a hypothesis for subsequent pharmacological evaluation. The translational relevance of these candidates may vary across prostate cancer disease states. Current treatment strategies differ substantially between hormone-sensitive prostate cancer and advanced disease states, including androgen receptor (AR)-positive castration-resistant prostate cancer, lineage-plastic prostate cancer, and neuroendocrine prostate cancer (Gillessen et al., 2025; Imamura et al., 2023). Because the present analyses evaluate genetic susceptibility rather than treatment response, metastatic progression, or molecular subtype-specific outcomes, these drug–target relationships should be considered hypothesis-generating candidates rather than direct therapeutic recommendations. Future studies integrating molecular subtyping and functional validation will be required to determine whether these targets may complement established approaches, including AR-directed therapy or radiotherapy. STRING associations linked several genetically prioritized candidate proteins to proteins associated with estradiol or etoposide. These connections nominate pharmacological relationships for subsequent experimental evaluation. Compared with bulk TWAS and GWAS-only approaches, our framework improves cellular resolution and provides additional biological context for interpreting susceptibility-associated signals. While previous prostate cancer TWAS studies leveraging bulk eQTL data have successfully nominated candidate eGenes, they are limited by tissue heterogeneity and inability to resolve cell-type-specific regulatory effects (Liu et al., 2022; Wu et al., 2019). In contrast, our integration of cell-resolved genetic regulation with tumor single-cell transcriptomics links genetically prioritized susceptibility signals to their transcriptional contexts within the prostate tumor microenvironment. This approach not only reveals novel mechanisms that would be obscured in bulk analyses but also provides a high-confidence set of functionally and clinically relevant targets for further exploration.

Despite its strengths, this study has several limitations that should be acknowledged. First, most genetic analyses were conducted in European ancestry populations, and the available scRNA-seq and sc-eQTL datasets had limited sample sizes. These factors may affect generalizability and the stability of associations, particularly in rare immune populations, highlighting the need for validation in larger and more diverse cohorts. Second, the present analysis did not systematically distinguish exhausted, cytotoxic, regulatory, or proliferative T-cell states, nor did it comprehensively resolve macrophage and myeloid phenotypes. Future studies integrating larger single-cell datasets with deeper immune-state annotation will be valuable for determining whether the prioritized genetic signals preferentially map to specific functional immune states. Third, although MR and colocalization strengthen causal inference by integrating genetically predicted expression with shared-signal evidence, they do not establish cell-state-specific biological mechanisms or therapeutic efficacy. In particular, the cellular context captured by an eQTL reflects the setting in which genetic regulation was detected, whereas the corresponding gene may retain broader expression and biological functions across other cellular compartments. Future spatial-transcriptomic analyses and functional perturbation studies will be required to validate the cellular localization, biological mechanisms, and therapeutic relevance of prioritized candidates.

In conclusion, this study presents an integrative framework combining immune-cell-resolved eQTLs, MR, colocalization, and tumor single-cell transcriptomics to prioritize prostate cancer susceptibility genes and place them within their relevant cellular contexts. The resulting genetic and transcriptional landscape highlights candidate immune-related regulatory programs and pharmacological targets for subsequent mechanistic and translational investigation. Future integration with functional experiments, spatial transcriptomics, and subtype-specific prostate cancer datasets will further refine their biological and therapeutic relevance.

## 4. Materials and methods

### 4.1 Data sources

This study was conducted in accordance with the STROBE-MR guidelines to ensure methodological transparency and reproducibility (Skrivankova et al., 2021). To define genetic instruments specific to immune cell gene expression, we utilized cis-eQTL data from the OneK1K sc-eQTL dataset (https://onek1k.org), which profiles 14 distinct human immune cell types (Yazar et al., 2022). In addition, bulk cis-eQTL data were obtained from the eQTLGen Consortium (https://eqtlgen.org), which provides genetic and transcriptomic data from 31,684 individuals of predominantly European ancestry, identifying significant cis-eQTLs for 16,987 genes (Võsa et al., 2021). Genome-wide association summary statistics for prostate cancer were obtained from the PRACTICAL consortium (Prostate Cancer Association Group to Investigate Cancer Associated Alterations in the Genome) (Schumacher et al., 2018). For replication, we used GWAS summary statistics of prostate cancer from FinnGen release 12 (R12), with the endpoint code C3_PROSTATE_EXALLC. FinnGen integrates genetic data from Finnish biobanks with nationwide health registry records (https://www.finngen.fi/en) (Kurki et al., 2023). Detailed information on the GWAS datasets used in this study is included in ***Table S1***.

### 4.2 Instrument selection

To ensure robust instrument validity, cis-eQTL variants meeting a prespecified association threshold of *p* < 1.0 × 10^-5^ were used as candidate instruments (Ray et al., 2025). Linkage disequilibrium (LD) clumping was performed using the 1000 Genomes Project European reference panel, with an *r^2^* threshold of 0.001 and clumping windows of 10,000 kb for single-cell eQTL instruments and 1,000 kb for bulk-blood eQTL instruments (Gkatzionis et al., 2023). For harmonization between exposure and outcome datasets, we aligned effect alleles and excluded ambiguous or strand-incompatible single nucleotide polymorphisms (SNPs). Palindromic variants were retained only when allele frequency permitted unambiguous interpretation. Following harmonization, SNPs associated with prostate cancer at *p* ≤ 1.0 × 10^-5^ in the outcome GWAS were excluded from the MR analysis. To prevent weak instrument bias, SNPs with *F*-statistics < 10 were excluded from the analysis (Pierce et al., 2011).

### 4.3 MR analysis

We conducted MR analyses to estimate the genetically predicted effects of gene expression, instrumented within individual immune-cell populations, on prostate cancer susceptibility. All analyses were performed using the TwoSampleMR R package (v0.6.6) (Hemani et al., 2018). For each eGene, the MR method was selected based on the number of available independent SNP instruments. When only a single SNP was available, the Wald ratio method was applied; for eGenes with two or more independent SNPs, the inverse-variance weighted (IVW) method was used instead (Burgess et al., 2023). Due to the limited number of SNP instruments, sensitivity analyses, including heterogeneity and pleiotropy tests, were not conducted. Multiple testing in the PRACTICAL discovery MR analyses was controlled using the Benjamini–Hochberg (BH) procedure. For the sc-eQTL analyses, correction was applied jointly across all eGene–cell-type MR tests spanning 14 immune cell types. For the bulk-blood eQTL analysis, correction was performed separately across all gene-level MR tests. In both analyses, the *p*-values from the Wald ratio or inverse-variance weighted estimates were adjusted before candidate selection, and associations with false discovery rate (FDR) < 0.05 were considered statistically significant. After BH correction, entries with unresolved gene symbols were excluded without recalculating adjusted *p*-values.

### 4.4 Weighted correlation analysis of MR effect estimates

To assess the concordance of MR effect estimates across eQTL contexts, we calculated weighted Pearson correlations after matching genes by identifier. Cell-type–bulk comparisons were weighted by the inverse of the mean variance of the two MR estimates, whereas comparisons between cell types used the geometric mean of their inverse-variance weights. A small constant (10^-8^) was added to stabilize the weights. Each comparison required at least 20 overlapping genes with valid estimates and weights. For cell-type–bulk comparisons, two-sided *p*-values were obtained from weighted linear regression slope tests and were reported without multiple-testing correction.

### 4.5 Bayesian colocalization analysis

To assess whether gene expression and prostate cancer susceptibility share a causal variant, we performed Bayesian colocalization analysis using the coloc R package (v5.2.3) (Giambartolomei et al., 2014). Analyses were restricted to eGene–cell-type associations meeting FDR < 0.05 in the discovery MR analysis. For each association, eQTL and prostate cancer GWAS summary statistics were matched by SNP identifier, duplicate SNP records were removed, and allele alignment was checked. Gene expression was modeled as a quantitative trait and prostate cancer as a case–control trait. Posterior probabilities for five competing hypotheses were calculated using coloc.abf, with prior probabilities of p_1_ = 1 × 10^-4^, p_2_ = 1 × 10^-4^, and p_12_ = 1 × 10^-5^. These represent the prior probabilities that a variant is associated with gene expression, prostate cancer susceptibility, or both traits, respectively. We focused on PP.H4, the posterior probability that both traits are associated and share a single causal variant. eGene–cell-type associations with PP.H4 ≥ 0.50 were considered to show colocalization evidence, with PP.H4 ≥ 0.70 indicating strong support.

### 4.6 Protein association network construction and topological analysis

Unique eGenes with colocalization evidence (PP.H4 ≥ 0.50) in at least one immune-cell eQTL context were queried against the STRING database (https://string-db.org) to retrieve functional associations among their encoded proteins (Szklarczyk et al., 2023). Gene identifiers were mapped to STRING protein identifiers, and the returned protein identifiers and preferred names were retained for network representation. Associations with a combined score ≥ 0.15 were retained. The resulting network was visualized in Cytoscape (v3.10.2), and node degree was used to characterize protein connectivity.

### 4.7 Cross-cohort replication and meta-analysis

To further assess the consistency of MR associations across cohorts, we combined estimates from the PRACTICAL discovery cohort and the FinnGen replication cohort using random-effects meta-analysis implemented in the metafor R package (v4.8) (Viechtbauer, 2010). Between-study variance was estimated using restricted maximum likelihood (REML). eGene–cell-type associations with meta-analysis *p* < 0.05 were considered nominally significant in the combined analysis. For visualization of cross-cohort prioritized associations, we displayed associations with strong colocalization evidence in the PRACTICAL cohort (PP.H4 ≥ 0.70), available FinnGen replication estimates, and significant random-effects meta-analysis results (*p* < 0.05).

### 4.8 Functional enrichment and network module analysis

To characterize the biological functions of the broader MR-prioritized candidate set, we performed functional enrichment analysis using Metascape (http://metascape.org) (Zhou et al., 2019). The reported enrichment results included GO Biological Processes, KEGG Pathway, Reactome Gene Sets, WikiPathways, Canonical Pathways, and Comprehensive Resource of Mammalian Protein Complexes (CORUM). Densely connected modules within the protein association network were identified using the MCODE algorithm, followed by module-specific functional enrichment analysis (Bader & Hogue, 2003). All visualizations were generated using ggplot2 (v3.5.2) in R and Cytoscape (v3.10.2).

### 4.9 The scRNA-seq data pre-processing and annotation

To contextualize genetically prioritized genes within the prostate tumor microenvironment, we reanalyzed the publicly available scRNA-seq dataset GSE137829 (https://www.ncbi.nlm.nih.gov/geo/), generated from prostate needle-biopsy specimens from six patients with castration-resistant prostate cancer (CRPC) (Dong et al., 2020). Expression matrices were processed using Seurat R package (v5.1.0) (Hao et al., 2024). Within each sample, cells with fewer than 300 detected genes were initially excluded, followed by removal of genes detected in fewer than three retained cells. After merging samples, additional cell-level quality control retained cells with >200 and <10,000 detected genes, >250 and <50,000 total RNA counts, <25% mitochondrial gene counts, and <10% hemoglobin gene counts. These steps retained 25,574 cells for subsequent clustering. Data were normalized and variance-stabilized using SCTransform, followed by principal component analysis (PCA) (Choudhary & Satija, 2022; Hafemeister & Satija, 2019). To correct for batch effects, we applied the Harmony algorithm, and unsupervised clustering was performed using the FindClusters function (resolution = 0.4) (Korsunsky et al., 2019). Cell type annotation was based on canonical markers and the unique expression profiles of cluster-specific genes. Subsequently, we extracted T cells, B cells, and plasma cells because these lymphocyte populations overlapped with major cellular contexts represented in the sc-eQTL analysis. We reclustered these cells at a higher resolution (resolution = 1) to examine whether prioritized genes showed differential transcriptional representation across the major lymphocyte populations within the prostate tumor microenvironment.

### 4.10 Drug repurposing

To explore pharmacological connections involving genetically prioritized prostate cancer susceptibility genes, we queried DrugBank (https://go.drugbank.com) using their gene symbols and retrieved associated compounds and drug–protein records (Knox et al., 2024). Separately, we compiled DrugBank records for drugs and diagnostic agents associated with prostate cancer, together with their associated proteins. These records were interpreted according to the annotated protein roles, including pharmacological targets, enzymes, transporters, and carriers. We then used STRING (https://string-db.org) to examine associations between the genetically prioritized candidate proteins and the proteins associated with these agents, retaining associations with a combined score ≥ 0.40 (medium confidence) (Szklarczyk et al., 2023). This analysis was designed to generate pharmacological hypotheses. A DrugBank association may reflect a pharmacodynamic or pharmacokinetic relationship, while a STRING association may reflect a functional relationship rather than direct physical binding. Neither relationship alone establishes that a compound modulates the genetically prioritized protein in a therapeutically beneficial direction.

## Supporting information

STROBE-MR-checklist

MDAR checklist

## Acknowledgements

The authors would like to thank the contributors of the PRACTICAL consortium, FinnGen consortium, eQTLGen consortium, and the OneK1K project for providing access to invaluable GWAS and eQTL datasets that made this study possible. We also acknowledge the GEO database (GSE137829) for the availability of prostate cancer single-cell RNA-seq data. We are grateful to the developers of TwoSampleMR, coloc, Seurat, Metascape, and STRING for the open-source tools that supported our analysis. Special thanks to Yuze Mi from Wenzhou Medical University for his assistance in detail refinement and visualization optimization during the manuscript drafting stage.

## Additional information

### Author contributions

Yanggang Hong: Conceptualization, Investigation, Formal analysis, Data curation, Validation, Visualization, Writing - original draft, Writing - review & editing, Supervision, Project administration. Yi Wang: Writing - original draft. Yirong Wang: Writing - original draft. Feng Chen: Validation. Jiajun Li: Writing - review & editing.

### Ethics

The analyses for this study were based on publicly available datasets, and no additional ethics approval or consent was needed.

## Additional files

### Supplementary files

Supplementary File 1. Supplementary figures.

Supplementary File 2. Supplementary tables.

STROBE-MR-checklist

MDAR checklist

## Data availability

The datasets analyzed in this study were obtained from publicly accessible repositories. Single-cell eQTL data were retrieved from the OneK1K project (https://onek1k.org), and bulk eQTL data were sourced from the eQTLGen Consortium (https://www.eqtlgen.org). Prostate cancer GWAS summary statistics were obtained from the PRACTICAL consortium and the FinnGen consortium (https://www.finngen.fi/en). Single-cell RNA sequencing data were downloaded from the GEO database under accession number GSE137829. All other relevant data are included in the article and its supplementary files. All analyses were performed using R (v4.3.1). The following R packages were used: TwoSampleMR (v0.6.6) for Mendelian randomization, coloc (v5.2.3) for Bayesian colocalization, Seurat (v5.1.0) for single-cell RNA-seq processing, Harmony (v1.0) for batch correction, metafor (v4.8) for meta-analysis, and ggplot2 (v3.5.2) for data visualization. Custom R scripts used in this study are available from the corresponding author Yanggang Hong upon reasonable request.

The following previously published datasets were used:

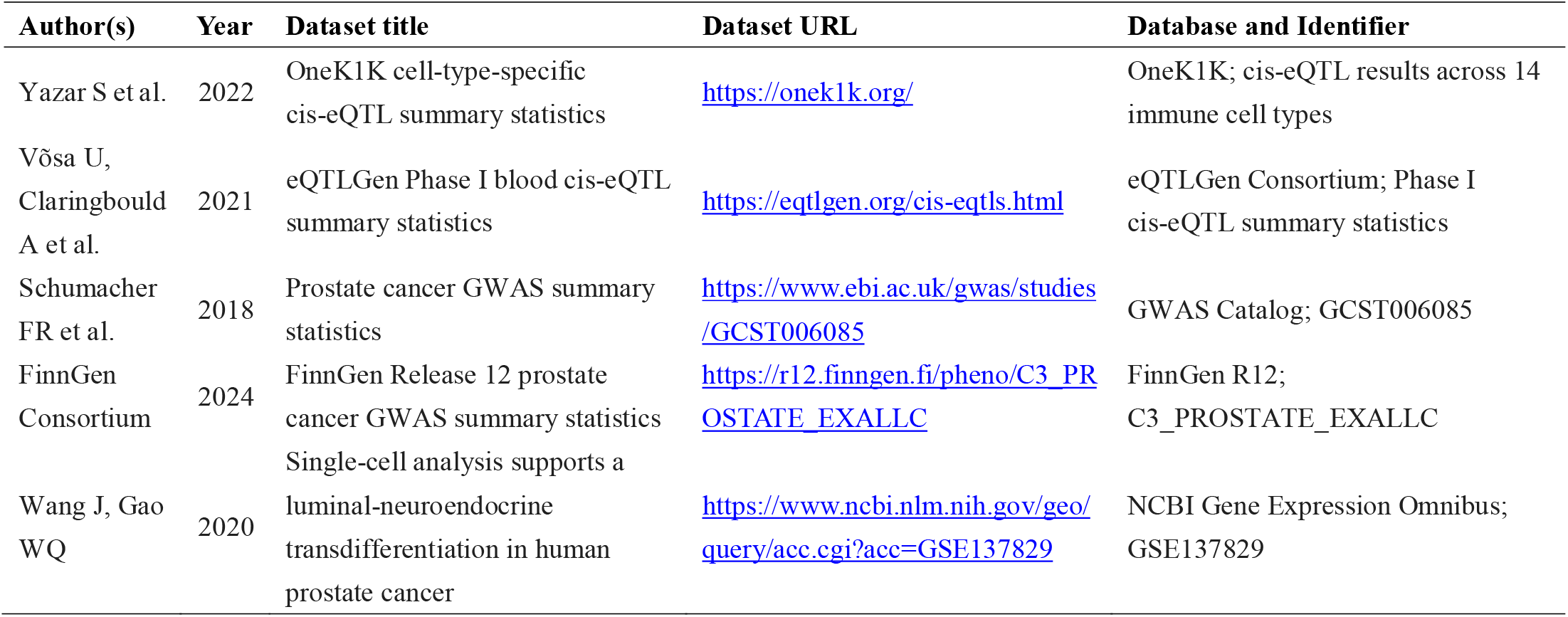

