## Supplementary File 1 for "Single-cell transcriptome-wide Mendelian randomization and colocalization analyses reveal immune-cell-specific mechanisms and actionable drug targets in prostate cancer"

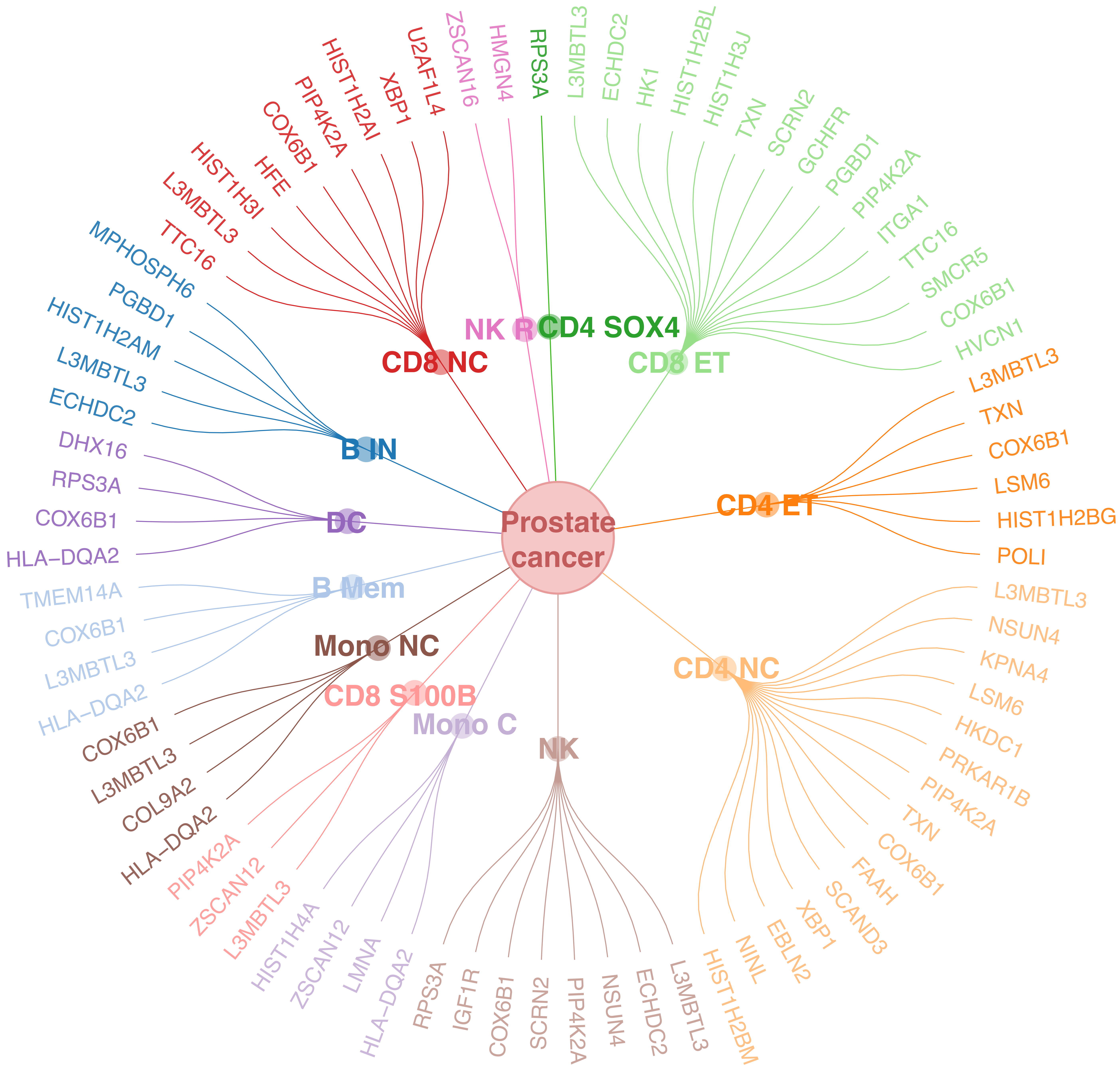

Prostate cancer
CD8 NC
U2AF1L4
XBP1
HIST1H2AI
PIP4K2A
COX6B1
HFE
HIST1H3I
L3MBTL3
TTC16
NK R
ZSCAN16
HMGN4
CD4 SOX4
RPS3A
CD8 ET
L3MBTL3
ECHDC2
HK1
HIST1H2BL
HIST1H3J
TXN
SCRN2
GCHFR
PGBD1
PIP4K2A
ITGA1
TTC16
SMCR5
COX6B1
HVCN1
CD4 ET
L3MBTL3
TXN
COX6B1
LSM6
HIST1H2BG
POLI
CD4 NC
L3MBTL3
NSUN4
KPNA4
LSM6
HKDC1
PRKAR1B
PIP4K2A
TXN
COX6B1
FAAH
SCAND3
XBP1
EBLN2
NINL
HIST1H2BM
NK
RPS3A
IGF1R
COX6B1
SCRN2
PIP4K2A
NSUN4
ECHDC2
L3MBTL3
Mono C
HIST1H4A
ZSCAN12
LMNA
HLA−DQA2
CD8 S100B
PIP4K2A
ZSCAN12
L3MBTL3
Mono NC
COX6B1
L3MBTL3
COL9A2
HLA−DQA2
B Mem
TMEM14A
COX6B1
L3MBTL3
HLA−DQA2
DC
DHX16
RPS3A
COX6B1
HLA−DQA2
B IN
MPHOSPH6
PGBD1
HIST1H2AM
L3MBTL3
ECHDC2

**Supplementary Figure S1.** Circular diagram showing the immune-cell-resolved distribution of colocalized eGene–cell-type associations prioritized by Bayesian colocalization (PP.H4 ≥ 0.50).

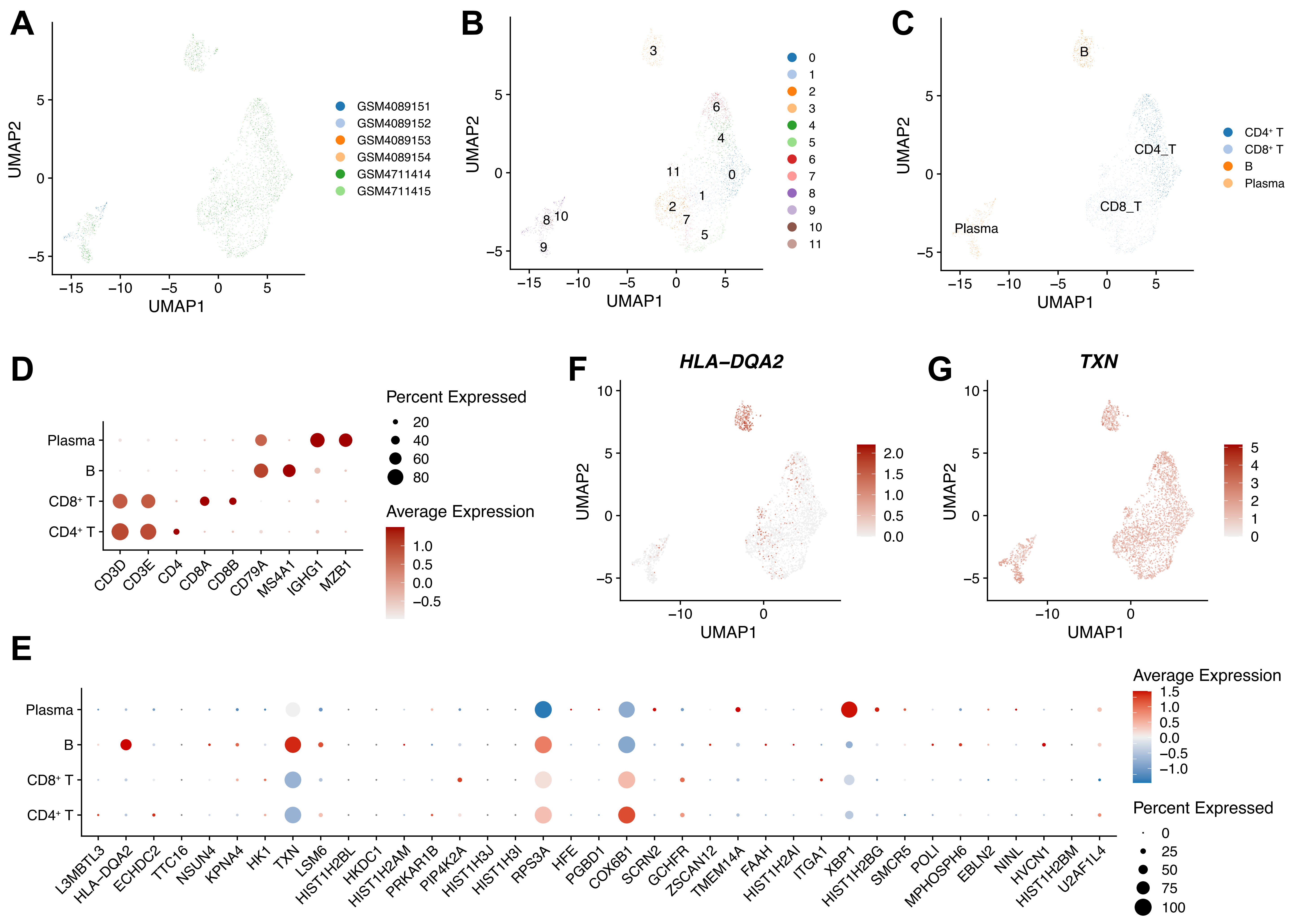
A
UMAP1
UMAP2
GSM4089151
GSM4089152
GSM4089153
GSM4089154
GSM4711414
GSM4711415
B
0 1 2 3 4 5 6 7 8 9 10 11
C
CD4⁺ T
CD8⁺ T
B
Plasma
CD4_T
CD8_T
D
Percent Expressed
20
40
60
80
Average Expression
1.0
0.5
0.0
−0.5
Plasma
B
CD8⁺ T
CD4⁺ T
CD3D CD3E CD4 CD8A CD8B CD79A MS4A1 IGHG1 MZB1
F
HLA−DQA2
G
TXN
E
Average Expression
1.5
1.0
0.5
0.0
−0.5
−1.0
Percent Expressed
0
25
50
75
100
L3MBTL3 HLA−DQA2 ECHDC2 TTC16 NSUN4 KPNA4 HK1 TXN LSM6 HIST1H2BL HKDC1 HIST1H2AM PRKAR1B PIP4K2A HIST1H3J HIST1H3I RPS3A HFE PGBD1 COX6B1 SCRN2 GCHFR ZSCAN12 TMEM14A FAAH HIST1H2AI ITGA1 XBP1 HIST1H2BG SMCR5 POLI MPHOSPH6 EBLN2 NINL HVCN1 HIST1H2BM U2AF1L4

**Supplementary Figure S2.** Transcriptional distribution of genetically prioritized eGenes across major lymphocyte populations. (A–C) UMAP plots of reclustered T cells, B cells, and plasma cells, colored by sample identity, unsupervised cluster, and annotated cell type. (D) Dot plot of canonical markers used for cell-type annotation. (E) Expression patterns of genetically prioritized eGenes across lymphocyte populations. (F–G) Feature plots of representative eGenes (*HLA-DQA2* and *TXN*) across reclustered immune populations.
