## Supplementary material for "Single-cell transcriptome-wide Mendelian randomization and colocalization analyses reveal immune-cell-specific mechanisms and actionable drug targets in prostate cancer": STROBE-MR-checklist

### STROBE-MR checklist of recommended items to address in reports of Mendelian randomization studies[1 2]

| Item No. | Section | | Checklist item | Page No. | Relevant text from manuscript |
|---|---|---|---|---|---|
| 1 | **TITLE and ABSTRACT** | | Indicate Mendelian randomization (MR) as the study's design in the title and/or the abstract if that is a main purpose of the study | 1, 3 | Title identifies "Mendelian randomization"; Abstract describes integration of cis-eQTL, GWAS, MR and colocalization. |
| | **INTRODUCTION** | | | | |
| 2 | **Background** | | Explain the scientific background and rationale for the reported study. What is the exposure? Is a potential causal relationship between exposure and outcome plausible? Justify why MR is a helpful method to address the study question | 4–5 | Introduction, paragraphs 1–4: prostate cancer burden, immune regulation, limitations of bulk eQTLs and rationale for MR. |
| 3 | **Objectives** | | State specific objectives clearly, including pre-specified causal hypotheses (if any). State that MR is a method that, under specific assumptions, intends to estimate causal effects | 5 | Introduction, final paragraph: aims to prioritize immune-cell-resolved eGenes, evaluate colocalization, replication and biological context. |
| | **METHODS** | | | | |
| 4 | **Study design and data sources** | | Present key elements of the study design early in the article. Consider including a table listing sources of data for all phases of the study. For each data source contributing to the analysis, describe the following: | 5, 10 | Study design overview (Figure 1); Methods 4.1 and Table S1 describe data sources. |
| | | a) | Setting: Describe the study design and the underlying population, if possible. Describe the setting, locations, and relevant dates, including periods of recruitment, exposure, follow-up, and data collection, when available. | 2, 10, 12 | Methods 4.1 and 4.9: OneK1K, eQTLGen, PRACTICAL, FinnGen R12 and GEO GSE137829; mainly European-ancestry eQTL/GWAS resources. |
| | | b) | Participants: Give the eligibility criteria, and the sources and methods of selection of participants. Report the sample size, and whether any power or sample size calculations were carried out prior to the main analysis | 2, 10, 12 | Methods 4.1, 4.9 and Table S1: source cohorts and study-level sample information; six CRPC biopsy samples in GSE137829. No prospective power calculation reported. |
| | | c) | Describe measurement, quality control and selection of genetic variants | 10–11 | Methods 4.2–4.3: cis-eQTL $p < 1 \times 10^{-5}$ , LD clumping, allele harmonization, outcome-associated SNP exclusion and F-statistic filtering. |
| | | d) | For each exposure, outcome, and other relevant variables, describe methods of assessment and diagnostic criteria for diseases | 10, 12 | Methods 4.1, 4.3 and 4.9: genetically predicted gene expression as exposure and prostate cancer susceptibility as outcome; FinnGen endpoint C3_PROSTATE_EXALLC. |
| | | e) | Provide details of ethics committee approval and participant informed consent, if relevant | 1 | Ethics: analyses used publicly available datasets; no additional ethics approval or consent was required. |
| 5 | **Assumptions** | | Explicitly state the three core IV assumptions for the main analysis (relevance, independence and exclusion restriction) as well assumptions for any additional or | 10–11, 9 | Methods 4.2–4.3 describe instrument filtering; Discussion discusses MR/colocalization limitations. The |

| | | | | |
|---|---|---|---|---|
| | | sensitivity analysis | | three IV assumptions are not explicitly enumerated. |
| 6 | **Statistical methods: main analysis** | Describe statistical methods and statistics used | 10–12 | Methods 4.2–4.8: instrument selection, MR estimation, weighted correlation, colocalization, meta-analysis and enrichment. |
| | a) | Describe how quantitative variables were handled in the analyses (i.e., scale, units, model) | 10–11 | Methods 4.3: gene-expression effects were genetically instrumented and related to prostate cancer susceptibility; MR effects presented as odds ratios. |
| | b) | Describe how genetic variants were handled in the analyses and, if applicable, how their weights were selected | 10–11 | Methods 4.2–4.3: independent cis-eQTL instruments selected by LD clumping and harmonized; Wald ratio for one SNP and IVW for ≥2 SNPs. |
| | c) | Describe the MR estimator (e.g. two-stage least squares, Wald ratio) and related statistics. Detail the included covariates and, in case of two-sample MR, whether the same covariate set was used for adjustment in the two samples | 10–11 | Methods 4.3: Wald ratio or inverse-variance weighted estimator according to instrument count. Cohort-specific covariate adjustment is not detailed. |
| | d) | Explain how missing data were addressed | 10, 7 | Methods 4.2: incompatible and ambiguous SNPs excluded; Results 2.6: five associations lacked sufficient overlapping instruments for replication. |
| | e) | If applicable, indicate how multiple testing was addressed | 11 | Methods 4.3: Benjamini–Hochberg adjustment across 14 immune-cell analyses jointly and separately for bulk blood; FDR < 0.05. |
| 7 | **Assessment of assumptions** | Describe any methods or prior knowledge used to assess the assumptions or justify their validity | 10–11 | Methods 4.2–4.5: instrument strength (F ≥ 10), allele harmonization, outcome-SNP filtering and Bayesian colocalization. No formal pleiotropy testing. |
| 8 | **Sensitivity analyses and additional analyses** | Describe any sensitivity analyses or additional analyses performed (e.g. comparison of effect estimates from different approaches, independent replication, bias analytic techniques, validation of instruments, simulations) | 7, 11–12 | Results 2.6 and Methods 4.5, 4.7: colocalization, FinnGen replication and random-effects meta-analysis; conventional heterogeneity and pleiotropy sensitivity tests not performed. |
| 9 | **Software and pre-registration** | | | |
| | a) | Name statistical software and package(s), including version and settings used | 2, 10–12 | Data availability lists R 4.3.1; TwoSampleMR 0.6.6, coloc 5.2.3, Seurat 5.1.0, Harmony 1.0, metafor 4.8, ggplot2 3.5.2. |
| | b) | State whether the study protocol and details were pre-registered (as well as when and where) | Not reported | No preregistration information is stated in the manuscript. |
| | **RESULTS** | | | |
| 10 | **Descriptive data** | | | |
| | a) | Report the numbers of individuals at each stage of included studies and reasons for | 2, 6–7, 12 | Data source table and Methods 4.9 report six CRPC samples and 25,574 QC-retained cells; Results 2.1 |

| | | | | | |
|---|---|---|---|---|---|
| | | | exclusion. Consider use of a flow diagram | | report 413 single-cell and 64 bulk MR associations. |
| | | b) | Report summary statistics for phenotypic exposure(s), outcome(s), and other relevant variables (e.g. means, SDs, proportions) | 2, 6–7 | Table S1 provides GWAS cohort information; Results 2.1–2.6 summarize associations. Individual-level phenotypic descriptive statistics are not supplied. |
| | | c) | If the data sources include meta-analyses of previous studies, provide the assessments of heterogeneity across these studies | 7, 11–12 | Methods 4.7: random-effects meta-analysis (REML) across PRACTICAL and FinnGen; no separate heterogeneity summary of source GWAS meta-analyses reported. |
| | | d) | For two-sample MR:<br>i. Provide justification of the similarity of the genetic variant-exposure associations between the exposure and outcome samples<br>ii. Provide information on the number of individuals who overlap between the exposure and outcome studies | 9–12 | Discussion notes European-ancestry predominance and limited generalizability; methods identify independent data resources. Participant overlap is not quantified. |
| 11 | **Main results** | | | | |
| | | a) | Report the associations between genetic variant and exposure, and between genetic variant and outcome, preferably on an interpretable scale | 6, 11 | Results 2.1 and Tables S2–S3 report gene-level MR results; Methods 4.2–4.3 describe genetic instrument associations. |
| | | b) | Report MR estimates of the relationship between exposure and outcome, and the measures of uncertainty from the MR analysis, on an interpretable scale, such as odds ratio or relative risk per SD difference | 6–7, 11 | Results 2.1 and 2.6, Tables S2–S3 and S17: MR and meta-analysis effect estimates, with odds ratios and 95% CIs shown in Figure 5. |
| | | c) | If relevant, consider translating estimates of relative risk into absolute risk for a meaningful time period | Not applicable | Absolute risk conversion was not performed; analyses address genetically predicted susceptibility associations. |
| | | d) | Consider plots to visualize results (e.g. forest plot, scatterplot of associations between genetic variants and outcome versus between genetic variants and exposure) | 20, 23 | Figure 2A displays weighted correlation of MR estimates; Figure 5 displays cross-cohort forest plots with odds ratios and 95% CIs. |
| 12 | **Assessment of assumptions** | | | | |
| | | a) | Report the assessment of the validity of the assumptions | 9–11 | Methods 4.2–4.5 and Discussion: instrument strength filtering and colocalization; assumptions cannot be fully verified with the available analyses. |
| | | b) | Report any additional statistics (e.g., assessments of heterogeneity across genetic variants, such as $I^2$, Q statistic or E-value) | 11–12 | Methods 4.3 notes heterogeneity and pleiotropy tests were not conducted owing to limited instrument counts. |
| 13 | **Sensitivity analyses and additional analyses** | | | | |

| | | | | |
|---|---|---|---|---|
| | | a) Report any sensitivity analyses to assess the robustness of the main results to violations of the assumptions | 9, 11–12 | Discussion describes residual causal-inference limitations; pleiotropy and heterogeneity sensitivity analyses were not performed. |
| | | b) Report results from other sensitivity analyses or additional analyses | 6–7, 11–12 | Results 2.2 and 2.6: colocalization and independent-cohort replication/meta-analysis. |
| | | c) Report any assessment of direction of causal relationship (e.g., bidirectional MR) | Not performed | No bidirectional MR or other formal assessment of causal direction was reported. |
| | | d) When relevant, report and compare with estimates from non-MR analyses | 6–7, 12 | Results 2.4–2.5: non-MR single-cell expression mapping provides biological context, rather than comparative effect estimates. |
| | | e) Consider additional plots to visualize results (e.g., leave-one-out analyses) | 23 | Figure 5 provides forest plots for replication/meta-analysis; leave-one-out plots were not generated. |
| | **DISCUSSION** | | | |
| 14 | **Key results** | Summarize key results with reference to study objectives | 8–9 | Discussion opening and Conclusion summarize immune-specific associations, colocalization, replication and therapeutic hypotheses. |
| 15 | **Limitations** | Discuss limitations of the study, taking into account the validity of the IV assumptions, other sources of potential bias, and imprecision. Discuss both direction and magnitude of any potential bias and any efforts to address them | 9 | Discussion limitations paragraph: ancestry, single-cell sample size, cellular resolution, MR assumptions and need for experimental validation. |
| 16 | **Interpretation** | | | |
| | | a) Meaning: Give a cautious overall interpretation of results in the context of their limitations and in comparison with other studies | 8–9 | Discussion evaluates results against prior prostate cancer TWAS and immune biology and interprets targets cautiously. |
| | | b) Mechanism: Discuss underlying biological mechanisms that could drive a potential causal relationship between the investigated exposure and the outcome, and whether the gene-environment equivalence assumption is reasonable. Use causal language carefully, clarifying that IV estimates may provide causal effects only under certain assumptions | 8–9 | Discussion: HLA antigen presentation, TXN redox control and COX6B1 mitochondrial metabolism are discussed as hypotheses, not established causal mechanisms. |
| | | c) Clinical relevance: Discuss whether the results have clinical or public policy relevance, and to what extent they inform effect sizes of possible interventions | 8–9 | Discussion: drug–target findings are hypothesis-generating and do not establish therapeutic efficacy. |
| 17 | **Generalizability** | Discuss the generalizability of the study results (a) to other populations, (b) across other exposure periods/timings, and (c) across other levels of exposure | 9 | Discussion limitations: predominant European ancestry, small scRNA-seq/sc-eQTL datasets, and limits of extrapolating to disease states and cellular contexts. |
| | **OTHER INFORMATION** | | | |
| 18 | **Funding** | Describe sources of funding and the role of funders in the present study and, if applicable, sources of funding for the databases and original study or studies on | 1 | Funding: "The authors declare that there was no funding |

| | | | | |
|---|---|---|---|---|
| | | which the present study is based | | for this work.” |
| 19 | **Data and data sharing** | Provide the data used to perform all analyses or report where and how the data can be accessed, and reference these sources in the article. Provide the statistical code needed to reproduce the results in the article, or report whether the code is publicly accessible and if so, where | 2 | Data availability: OneK1K, eQTLGen, PRACTICAL, FinnGen and GEO GSE137829; R code available from the corresponding author upon reasonable request. |
| 20 | **Conflicts of Interest** | All authors should declare all potential conflicts of interest | 1 | Competing interest: “The authors declare that no competing interests exist.” |

This checklist is copyrighted by the Equator Network under the Creative Commons Attribution 3.0 Unported (CC BY 3.0) license.

1. Skrivankova VW, Richmond RC, Woolf BAR, Yarmolinsky J, Davies NM, Swanson SA, et al. Strengthening the Reporting of Observational Studies in Epidemiology using Mendelian Randomization (STROBE-MR) Statement. JAMA. 2021;under review.

2. Skrivankova VW, Richmond RC, Woolf BAR, Davies NM, Swanson SA, VanderWeele TJ, et al. Strengthening the Reporting of Observational Studies in Epidemiology using Mendelian Randomisation (STROBE-MR): Explanation and Elaboration. BMJ. 2021;375:n2233.
